# Vision Language Models Fail to Reliably Detect Acute Myeloid Leukemia in Bone Marrow Smears

**DOI:** 10.64898/2026.08.19.26359329

**Authors:** Freya Schulze, Chiara Löffler, Martina Radoynova, Susann Winter, Christoph Röllig, Katja Sockel, Frank Kroschinsky, Martin Bornhäuser, Jan Moritz Middeke, Jakob Nikolas Kather, Jan-Niklas Eckardt, Narmin Ghaffari Laleh

## Abstract

Hematologic diagnostics and especially cytomorphologic assessment are time-intensive and require high levels of expertise. Vision Language Models (VLM) show promise in medical image analysis in radiology and histopathology, while an evaluation on detecting acute myeloid leukemia (AML) is lacking. Our goal was to evaluate three Vision Language Models regarding their diagnostic accuracy and safety in clinical decision support in detecting AML from digitized bone marrow smears (BMS). Whole slide images were obtained from bone marrow smears of 50 AML patients and 50 bone marrow donors. Ten representative fields of view per sample were extracted manually. Three VLMs were used, two of which are considered generalist models (Qwen3.5-397B-A17B-FP8, GLM-4.6V-FP8), while the other one is a medically adapted model (Medgemma-27b-it). All models performed zero-shot analysis using two prompting strategies: First, a context-rich prompt requesting reporting of WHO/FAB diagnostic criteria in a structured manner, and secondly a minimal prompt without specific hematologic context.

Overall diagnostic accuracy was poor for all models as they exhibited the overwhelming tendency to classify most samples as leukemic: With context-rich prompts, GLM4.6 identified 90% of leukemic samples while also labeling 92% of bone marrow donors as AML. The medical specialist model MedGemma-27b showed similar failure, misclassifying 86% of healthy donors and correctly detecting AML in only 66% of cases. Qwen3.5 performed best under detailed prompting, achieving a specificity of 0.26 and accuracy of 0.51. Accuracy of all models improved with context-free prompts (accuracies range 0.47-0.79), yet they still lacked the ability to correctly distinguish between leukemia and healthy bone marrow. Qwen3.5 was the only model to maintain meaningful specificity (0.64) and correctly identified 94% of AML, yielding an overall accuracy of 0.79. Morphologic feature-level agreement with human expert reports was poor across all models, indicating poor recognition of cell-level morphologies.

This failure is likely driven by the fact that pathology imaging archives are vastly scraped during model training while hematological samples are not as widely available and therefore, hematology is an out-of-bounds use-case for these models, rendering them currently unsuitable for clinical decision support in hematology.

## Main

Cytomorphologic analysis of bone marrow smears remains a cornerstone of hematologic diagnostics, particularly for classifying acute myeloid leukemia (AML), an aggressive and biologically heterogeneous disease requiring time-sensitive, precise diagnostics.^1^ Despite advances in molecular profiling and flow cytometry, microscopic analysis remains essential per WHO criteria.^2^ However, cytomorphologic assessment is time-intensive, requires years of experience, and is prone to inter-observer variability.^3^

Machine learning approaches have shown promise in AML diagnostics, spanning cytomorphologic classification, risk stratification, and treatment prediction.^4^ However, these efforts have largely relied on task-specific supervised models trained on annotated datasets. Vision Language Models (VLMs) represent a different paradigm: rather than learning from disease-specific labels, they perform zero-shot analysis, i.e., classification without task-specific examples, leveraging broad visual and linguistic knowledge from large-scale pretraining.^5^

VLMs have emerged as promising tools for automated image analysis across clinical disciplines.^6,7^ In radiology and histopathology, VLMs have moved from proof-of-concept to compelling applications including structured radiology report generation,^8,9^ tumor subtyping,^10^ and cancer grading.^11^

Cytomorphologic analysis of bone marrow smears poses unique challenges: staining variability, heterogeneous cellular composition, and a near-absence of hematology-specific training data.^12^ While a recent study benchmarked VLMs on peripheral blood and bone marrow cell classification,^13^ a systematic evaluation of state-of-the-art (SOTA) VLMs specifically on their ability to distinguish AML from healthy bone marrow at the patient level has not been reported. Here, we benchmark three SOTA open-source VLMs on this task in a zero-shot setting, assessing their readiness and safety for hematologic diagnostics.

Newly diagnosed AML patients were identified from the multicenter registry of the German Study Alliance Leukemia (NCT03188874). The study was approved by the Institutional Review Board of the Technical University Dresden. All patients and donors provided informed consent according to the revised Declaration of Helsinki. Whole slide images (WSIs) of bone marrow smears from 50 AML patients and 50 healthy bone marrow donors at University Hospital Dresden were matched 1:1 as a proof-of-concept cohort, enabling balanced sensitivity and specificity estimation. Given the labor-intensive manual extraction of regions of interest (ROIs) required per sample, this scale enabled rigorous zero-shot benchmarking across models and prompting strategies. Slides were scanned on a Pannoramic SCAN II (3DHISTECH; 20x objective, 1.6x C-mount adapter; 0.2035 µm/pixel). Ten representative ROIs per sample were manually extracted by two expert hematologists in SlideViewer (3DHISTECH) at 63x magnification (600 dots per inch; 3,591 × 2,870 pixels per ROI). AML was diagnosed according to WHO and International Consensus Classification criteria.^2,14^

Three state-of-the-art VLMs were evaluated: GLM-4.6V-FP8, Qwen3.5-397B-A17B-FP8, and MedGemma-27B-it. Models were selected to represent a range of architectures and adaptation strategies, with two large general-purpose VLMs (GLM-4.6, Qwen3.5) and one medically fine-tuned model (MedGemma-27B-it). All three were accessed via OpenAI-compatible programming interfaces (APIs). Models were prompted using two strategies: a context-rich prompt requesting structured cytomorphologic reporting per predefined diagnostic categories, and a context-free prompt asking only for the final classification (prompts in Supplementary Methods). Images were encoded as base64 inputs and submitted per patient, either individually or as a 10-ROI multi-image set. Inference used default model settings, with model thinking disabled where supported. We calculated overall accuracy, AML-positive sensitivity and specificity, macro-averaged sensitivity, specificity, F1, and a binary macro-F1 score for AML versus non-AML classification. For accuracy estimates, 95% Wilson confidence intervals quantified uncertainty, and one-sided binomial tests assessed whether accuracy exceeded chance-level baseline. Chance-level accuracy was 0.5 for the binary context-free task, and 1/7 for the seven-category context-rich task (Supplementary Methods). For Table 1, context-rich outputs were subsequently reduced to a binary AML versus non-AML outcome. Cohen’s kappa (κ) was calculated separately for each model against expert ground truth to assess feature-level agreement across 18 cytomorphologic categories (Figure 1A).

**Table 1.**
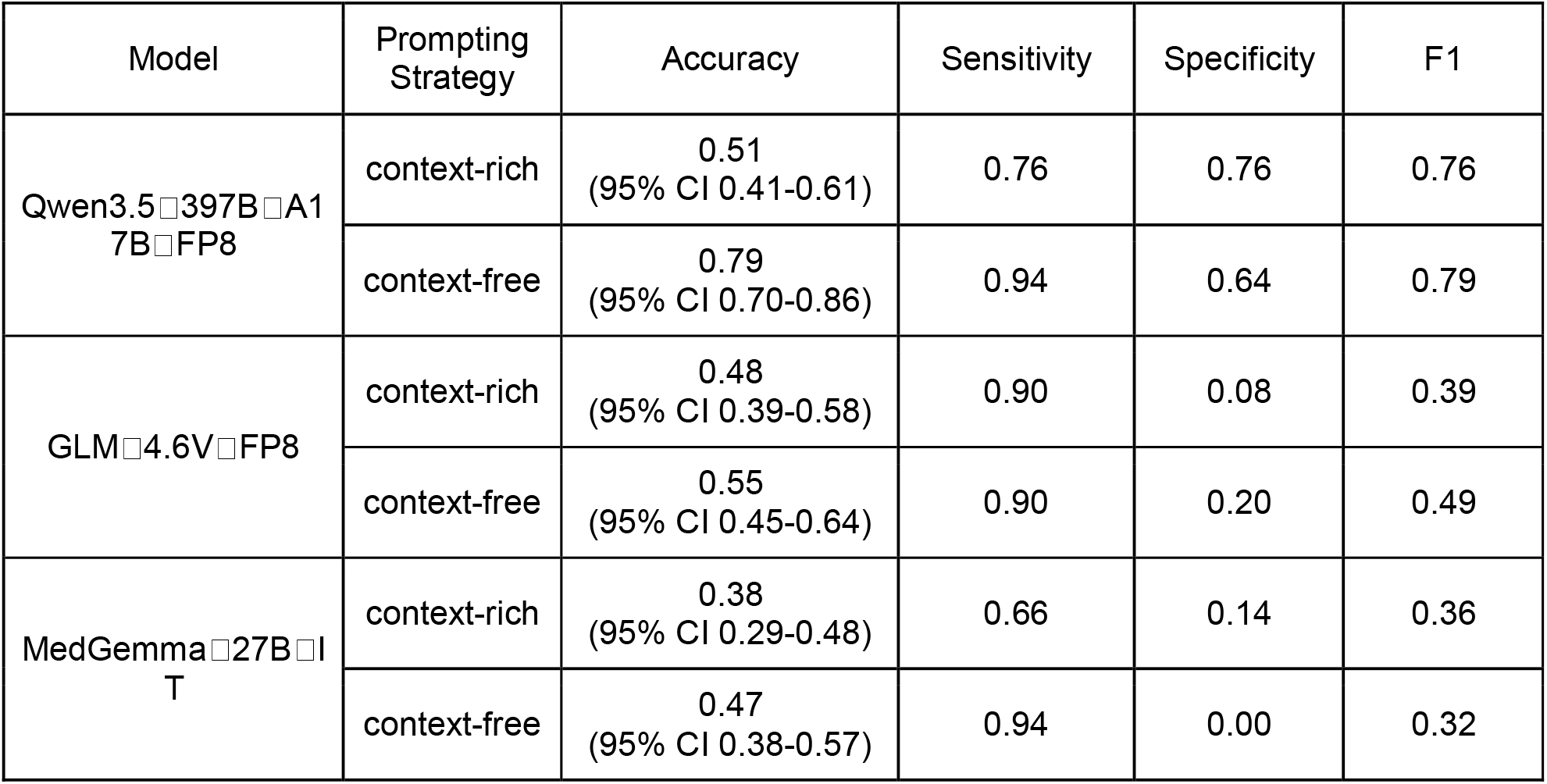
Summary of classification performance across vision–language models and prompting strategies.

**Figure 1.**
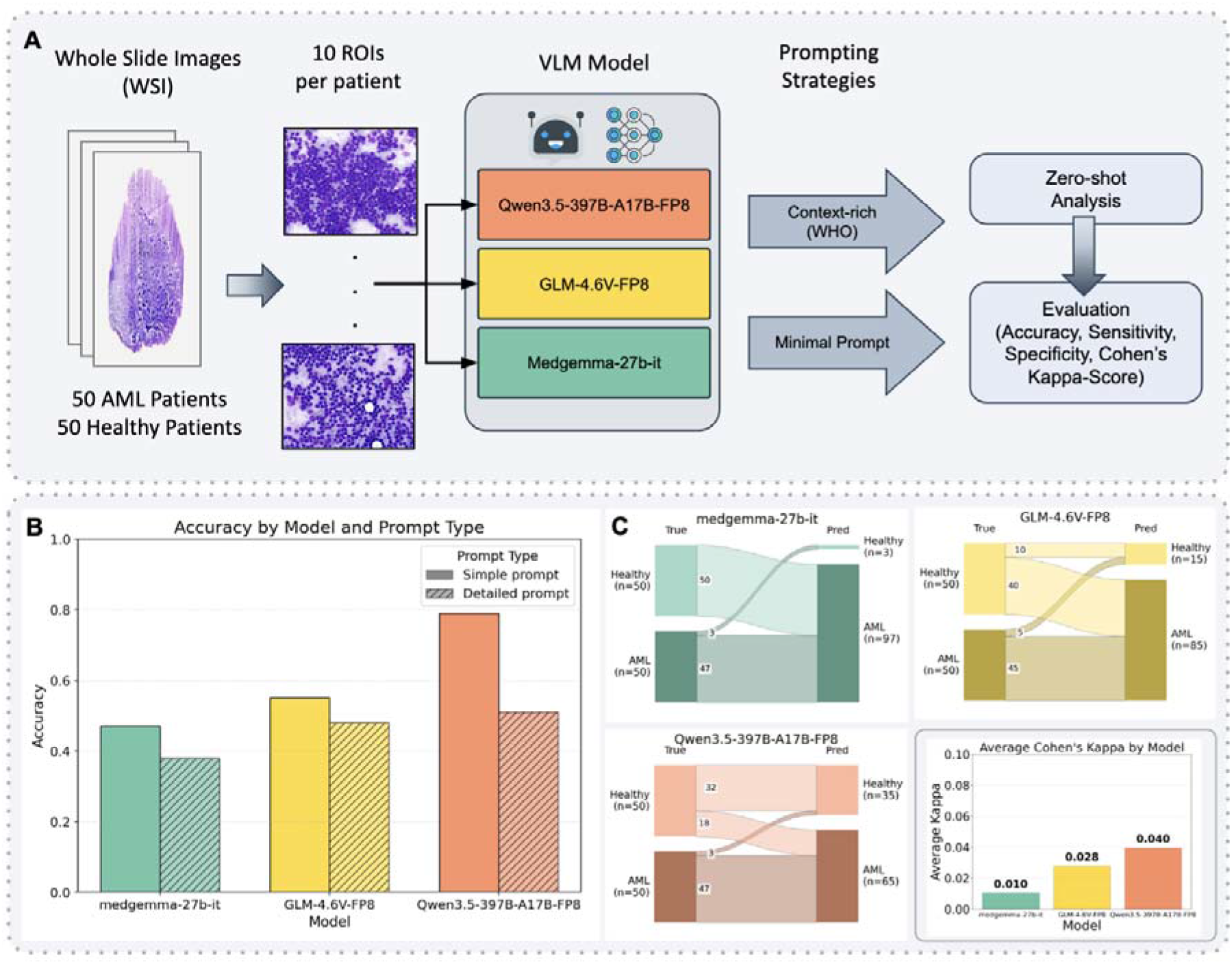
Study design, model evaluation, and classification performance for AML detection using vision–language models. (A) Workflow overview. Whole slide images (WSIs) from 50 acute myeloid leukemia (AML) patients and 50 healthy controls were sampled to generate 10 regions of interest (ROIs) per patient. These image patches were analyzed using three vision–language models (VLMs): Qwen3.5-397B-A17B-FP8, GLM-4.6V-FP8, and MedGemma-27B-it. Two prompting strategies were applied: context-rich prompts incorporating WHO information and minimal prompts. Model outputs were evaluated using accuracy, sensitivity, specificity, and Cohen’s kappa. (B) Classification accuracy across models and prompt types. (C) Prediction behavior under minimal prompting. AML, acute myeloid leukemia; ROI, region of interest; VLM, vision–language model; WSI, whole slide image; WHO, World Health Organization classification; Pred, predicted label; N-NMS, non-neoplastic marrow samples.

Under context-rich prompting, none of the three VLMs achieved clinically reliable discrimination between AML and healthy bone marrow. Qwen3.5 performed comparatively best, with sensitivity and specificity both at 0.76 and an accuracy of 0.51 (95% CI 0.41-0.61) (Figure 1B). However, its class-wise F1 score was markedly lower for healthy bone marrow (0.39) than for AML (0.76), indicating that even the most balanced sensitivity/specificity profile among the three models failed to reliably confirm healthy marrow, misclassifying 25% of healthy donors as leukemic. GLM-4.6 correctly identified most AML cases (sensitivity 0.90) but almost uniformly misclassified healthy marrow as leukemic (specificity 0.08), yielding an accuracy of 0.48 (95% CI 0.39-0.58), consistent with a positive bias reported in LLMs.^15^ MedGemma-27B-it showed lower sensitivity (0.66) with similarly poor specificity (0.14), resulting in the lowest overall accuracy of 0.38 (95% CI 0.29-0.48) and an F1 score of 0.36.

Under context-free prompting, Qwen3.5 stood out with the strongest diagnostic profile in this study, combining high AML sensitivity (0.94) with moderate specificity (0.64), yielding the highest overall accuracy (0.79, 95% CI 0.70-0.86) and the best class-wise F1 scores for both AML (0.82) and normal bone marrow (0.75). Even this result, however, corresponds to more than a third of healthy donors misclassified as leukemic, falling short of clinical acceptability. The remaining two models performed worse, with AML overcalling persisting throughout. GLM-4.6 maintained high sensitivity (0.90) but still failed to distinguish healthy from leukemic bone marrow (specificity 0.2, accuracy 0.55, 95% CI 0.45-0.64). MedGemma-27B-it achieved very high AML sensitivity (0.94) but completely failed to identify normal marrow, hallucinating blast cells and labeling every case leukemic (specificity 0.00, accuracy 0.47, 95% CI 0.38-0.57) (Figure 1C).

Feature-level concordance between model outputs and expert reports was poor across all models, regardless of prompting strategy. Mean Cohen’s κ was 0.025 (range 0.01 to 0.04) across 18 cytomorphologic categories under context-rich prompting, indicating poor recognition of cell-level morphologies. Dysplasia detection was absent across all three hematopoietic lineages, with mean κ near zero for granulopoiesis (κ = 0.001, range 0.000-0.022), megakaryopoiesis (κ = 0.016, range 0.001-0.021), and erythropoiesis (κ = 0.008, range 0.005-0.013). Agreement for Auer rods was very poor (mean κ = 0.01, range 0.0032-0.023), indicating that models did not reliably detect this feature.

Hallucination of AML-defining features was striking across models. In healthy donors, blast counts exceeding 20% were falsely reported by GLM□4.6 in 90% and by MedGemma□27B-it in 78% of cases, rendering both incapable of reliably excluding AML. MedGemma□27B-it reported blast counts exceeding 50% in 27 of 50 healthy donors. Qwen3.5 showed the lowest hallucination rate, yet still falsely reported blast cell counts at leukemic levels in 18% of healthy donors.

In this study, we demonstrate that SOTA VLMs fail to achieve clinically acceptable diagnostic performance in AML detection, with systematic hallucination of AML-defining features in healthy bone marrow donors representing the most critical safety concern.

The tendency to overcall AML, most pronounced under context-rich prompting, likely reflects a systematic bias rooted in the near-absence of hematology-specific training data in publicly available datasets. Models thus default to leukemic interpretations when confronted with unfamiliar morphology, rather than performing genuine cytomorphologic reasoning. Consistent with neuroradiology findings,^16^ our results suggest that domain complexity and limited training data representation are persistent barriers to clinical-grade VLM performance in specialized imaging. This aligns with a concurrent evaluation of VLMs on peripheral blood and bone marrow cytomorphology, which similarly reported near-random zero-shot accuracy and found that model-generated explanations frequently failed to correspond to the morphological features driving classification decisions, underscoring that current VLMs lack genuine morphological understanding.^13^

Qwen3.5 overcalled AML least across prompting strategies, consistent with reports that large general-purpose VLMs can match or surpass medically fine-tuned models,^17^ yet it still misclassified up to one in three healthy donors as leukemic. Conversely, MedGemma-27B-it, although medically adapted, performed worse than the generalist models.^18^ Importantly, MedGemma’s adaptation did not encompass hematological cytomorphology, suggesting that cross-domain medical fine-tuning does not substitute for disease-specific training data and may bias models toward overrepresented modalities.^19^

Superiority of context-free prompting may reflect anchoring bias: explicit cytomorphologic criteria may have steered models toward leukemic classifications regardless of visual input.^20^ In a real diagnostic setting, these hallucinations would translate directly into false-positive AML diagnoses, triggering further diagnostics, prolonged hospitalization, and physical and psychological harm to healthy individuals, rendering current VLMs not suitable for AML diagnostics.

The evaluation cohort was drawn from a single center (50 AML patients, 50 healthy donors), and performance may vary with differences in staining protocols, slide digitization, manually curated ROIs, or patient population across institutions. We restricted evaluation to three open-source, locally deployable VLMs to safeguard patient data security. Newer models have since superseded those tested, and our findings reflect a snapshot of VLM capability rather than long-term limits. Still, the consistent hallucination patterns across architecturally distinct models suggest that scaling alone is unlikely to resolve these domain-specific reasoning failures without dedicated hematology training data. Although two distinct prompting strategies were tested, the observed failures, including higher accuracy under minimal context, indicate that prompt design cannot substitute for morphological reasoning. Finally, all models were evaluated in a zero-shot setting without any hematology-specific fine-tuning or in-context learning, representing both the key novelty of this study and an inherent performance constraint.

VLMs are currently inadequate for leukemia diagnostics. Clinical deployment of VLMs in hematology will require hematology-specific adaptation on rigorously annotated datasets, alongside transparent hallucination reporting and prospective benchmarking against specialist performance.

## Data Availability

All data produced in the present study are available upon reasonable request to the authors

## Supplementary Methods

### Prompts

#### Context-free prompt

~~~
{
“role”: “system”,
“content”: “““
You are a hematologist specializing in bone marrow cytomorphology. Your task is to microscopically analyze 10 ROI images of a bone marrow smear.
Objective: Based on the image data, create a **structured cytomorphological overall report** as well as a **general morphological diagnostic category**.
“““
},
{
“role”: “user”, “content”: [
{
“type”: “text”,
“text”: f”““
Please analyze the following 10 bone marrow ROIs. Create a cytomorphological description and a diagnosis. Provide the final answer exclusively in German in the following JSON format:
{{
“gedanken”: “Summary of the overall cytomorphological findings and diagnostic considerations.”,
“Diagnose”: Brief diagnosis in 1 sentence,
}}
~~~

#### Context-rich prompt

~~~
{
“role”: “system”,
“content”: “““
You are a hematologist specializing in bone marrow cytomorphology. Your task is to microscopically analyze 10 ROI images of a bone marrow smear that have been provided.
Create a structured cytomorphological report based on the visible morphological features.
Important notes:
-Describe objectively what you see
-The answer must be provided exclusively in **German**
-Use **only** the specified categories for structured fields
-Submit your answer **only in the defined JSON format**
“““
},
{
“role”: “user”, “content”: [
{
“type”: “text”,
“text”: f”““
Please systematically analyze the following 10 bone marrow ROIs and evaluate:
-Quality and cell content of the smear
-Erythropoiesis, granulopoiesis, megakaryopoiesis (quantity and dysplasia)
-Lymphopoiesis, monopoiesis, plasma cells
-Blasts (quantity and morphology)
-Special morphological features
Output the final answer exclusively in the following JSON format:
{{
“Gedanken”: “Your cytomorphological assessment and diagnostic classification (free text)”,
“fragments”: “SELECT EXACT: Sufficient | minimal | absent | N/A”,
“cell_count”: “SELECT EXACT: hypocellular | normal | hypercellular | cannot be assessed | N/A”,
“erythropoiesis”: “SELECT EXACT: hypocellular | normocellular | hypercellular | absent | N/A”,
“erythropoiesis_dysplasia_signs”: “SELECT EXACT: no | <10% | >10% |
>50% | N/A”,
“granulopoiesis”: “SELECT EXACT: hypocellular | normocellular | hypercellular | absent | N/A”,
“granulopoiesis_dysplasia_signs”: “SELECT EXACT: no | <10% | >10% |
>50% | N/A”,
“megakaryopoiesis”: “SELECT EXACT: hypocellular | normocellular | hypercellular | absent | N/A”,
“megakaryopoiesis_dysplasia_signs”: “SELECT EXACT: no | <10% | >10% |
>50% | N/A”,
“lymphopoiesis”: “SELECT EXACT: decreased | normal | increased | atypical cells | absent | N/A”,
“monopoiesis”: “SELECT EXACT: decreased | normal | increased | absent | N/A”,
“plasma cells”: “SELECT EXACT: decreased | normal | increased | absent
| N/A”,
“blast_percentage”: “SELECT EXACT: <5 | 5-19 | >20 | >50 | N/A”, “blast_cytoplasm”: “SELECT EXACT: basophilic | eosinophilic |
nonspecific | sparse | N/A”,
“nucleoli”: “SELECT EXACT: yes | no | N/A”, “granules”: “SELECT EXACT: yes | no | N/A”,
“chromatin”: “SELECT EXACT: fine | dense | coarse | nonspecific” “auer rods”: “SELECT EXACT: yes | no | N/A”,
“special_findings”: “Free text: e.g., cup-shaped nuclei, atypical mitoses, nuclear shadows, Faggot cells, macrophages with hemophagocytosis, or ‘no special findings’”,
“diagnosis”: “SELECT EXACT: Acute myeloid leukemia | MDS | Myeloproliferative neoplasm | Lymphoproliferative neoplasm | Normal BM
| Punctio sicca | N/A”
}}
**IMPORTANT:**
-For structured fields, use the EXACT terms provided (including uppercase/lowercase letters and special characters)
-If information is missing, use “N/A”
-For normal/healthy findings, select “Normal BM” as the diagnosis
-Ensure correct spelling: “Adequate” (capital A), “<10%” (with %), “N/A” (sometimes without a period)
“““
}
# Your images go here
]
}
~~~

